# How long and effective does a mask protect you from an infected person who emits corona virus-laden particles: by implementing physics-based modeling

**DOI:** 10.1101/2022.07.05.22277221

**Authors:** Flora Bahrami, Till Batt, Seraina Schudel, Simon Annaheim, Weidong He, Jing Wang, René M. Rossi, Thijs Defraeye

## Abstract

SARS-CoV-2 spreads via droplets, aerosols, and smear infection. From the beginning of the COVID-19 pandemic, using a facemask in different locations was recommended to slow down the spread of the virus. To evaluate facemasks’ performance, masks’ filtration efficiency is tested for a range of particle sizes. Although such tests quantify the blockage of the mask for a range of particle sizes, the test does not quantify the cumulative amount of virus-laden particles inhaled or exhaled by its wearer. In this study, we quantify the accumulated viruses that the healthy person inhales as a function of time, activity level, type of mask, and room condition using a physics-based model. We considered different types of masks, such as surgical masks and filtering facepieces (FFPs), and different characteristics of public places such as office rooms, buses, trains, and airplanes. To do such quantification, we implemented a physics-based model of the mask. Our results confirm the importance of both people wearing a mask compared to when only one wears the mask. The protection time before the healthy wearer has an infection risk of 50% reduces by 80% if only one wears the facemask instead of both people. The protection time is further reduced if the infected person starts to cough or increases the activity level by 85% and 99%, respectively. Results show the leakage of the mask can considerably affect the performance of the mask. For the surgical mask, the apparent filtration efficiency reduces by 75% with such a leakage, which cannot provide sufficient protection despite the high filtration efficiency of the mask. The facemask model presented provides key input in order to evaluate the protection of masks for different conditions in public places. The physics-based model of the facemask is provided as an online application.

## 1 Introduction

The first cases of coronavirus disease 2019 (Covid-19) were reported in December 2019 [1]. A worldwide pandemic followed the spread of the virus; on March 11, 2020, World Health Organization (WHO) characterized the Covid-19 outbreak as a pandemic [2]. By the beginning of July 2022, according to WHO coronavirus dashboard data, more than 540 million cases of infection have been confirmed globally, and more than 6.3 million people passed away as a result of Covid-19. Throughout the pandemic, the usage of the mask was controversial. In January 2020, WHO announced that the usage of the medical mask is not required as there is no evidence to protect healthy people. In April 2020, WHO changed the announcement and recommended wearing a medical mask for healthy people who care for an infected person. WHO changed the guidelines for using facemasks by advising to wear them in public when social distancing was not possible in June 2020. Since the first Covid-19 case report, the importance of wearing a facemask has shown its value in protecting people against this disease [3].

A substantial amount of research has been done on how different types of face masks protect the wearers from infection. Standardized mask performance analyses include experiments with controlled laboratory conditions. Here, mask filtration efficiency and pressure (breathing) resistance are measured [4–8]. Several standards were already available for different types of facemasks, such as FFP masks (EN 149:2009-08) or surgical facemasks (EN 14683:2019-10). Additionally, different experimental setups have been designed to mimic sneezing or coughing and to investigate their effect on filtration efficiency [9–12]. Experimental filtration efficiency tests give a good indication to evaluate and compare the performance of different mask types. Nevertheless, these tests do not provide information on users’ accumulation of exhaled or inhaled virus-laden particles/aerosols. This cumulative amount depends on the wearer’s breathing rate and the concentration of virus-laden particles in the wearer’s environment. Such information is challenging to measure experimentally. Therefore, researchers have explored mathematical modeling to monitor the fate of exhaled aerosols by an infected person to overcome this hurdle. Heretofore, several studies developed CFD models to predict the aerosol disposition in lungs, masks, and environments [13–17]. These studies provide key information on the risk for a healthy person. Yet, these studies do not explore the effect of different conditions such as environment, activity levels, and different types of masks. Besides the additional information provided by these models, they still do not quantify the risk of infection for people in the environment, as no integration over time is considered. Only a few physics-based models quantify the risk for a healthy person in different scenarios [18], [19].

This study aimed to quantify how long different mask types, such as surgical, FFP, and community masks, protect the wearer in four different environments at five different activity levels. The studied community mask in this study was a textile mask compatible with Swiss rule (SNR 30000). The surgical, FFP, and community masks were studied in different environments such as an office room, train, bus, and airplane. We developed a physics-based computational replica of the facemasks that simulates the inflow and outflow of virus-laden particles for healthy and infected wearers. The model accounts for two people: one infected with the Covid-19 virus, and the other one is healthy. Different types of activities were simulated relevant for various activities, from sitting to more vigorous activities such as running. The effect of speaking and coughing during contact time with a healthy person was also included in the model. Using this model, we quantified the protection time of standard masks for a healthy person in different environments.

## 2 Background

We sketch the main characteristics of mask protection against virus-laden particles. These traits will define the testing environment we later used for the simulations.

### 2.1 Breathing/Speaking/Coughing

Humans breathe continuously, where the duration of inhalation is usually shorter than exhalation. The breathing rate and its pattern depend on the activity level. We exhale endogenously generated particles during breathing, and when we start to speak or cough, larger droplets in higher quantities can be generated [20–22]. Furthermore, the measured median diameter of the droplets we exhale varies for different activities resulting in a whole droplet size distribution range of 0.1-1,000,000 nm [23]. Due to the gravitational forces, the droplet > 5µm usually settles rather fast in the exhaled air, i.e., within a meter [24,25]. In addition, the size of the droplets and aerosols can decrease very rapidly due to evaporation in the environment. Most of the emitted aerosols during breathing, speaking, and coughing have a diameter between 100 to 2000 nm [21]. The SARS-CoV-2 virus itself has a diameter size ranging from 65–125 nm [26], so several viruses can be contained in a single droplet or aerosol. The highest risk of contamination from an infected person for people at a distance comes from the smaller virus-laden particles, so aerosols [27]. The smallest particles can travel farther or can circulate indoors [28].

### 2.2 Mask and Filtration

Facemasks filter out aerosols and particles from the air. Several types of masks and respirators are now being used to protect from aerosol transmissible diseases, such as community masks, surgical masks, particle filtering half masks, and disposable filtering facepieces (FFP). Masks are filters that stop the particles by different modes of action: interception, inertial impaction, electrostatic deposition, and diffusion [29]. For sub-micron-sized aerosols generated by breathing, masks also rely on electrostatic deposition. This filtration by each of these phenomena strongly depends on the filter, particles’ size and characteristics, and additional parameters like air flow rate and the filter properties of the mask. The resulting combination of these filtration effects is quantified for each particle size in a single metric, namely the particles filtration efficiency. The efficiency (*FE*) of the filtering device is defined as:

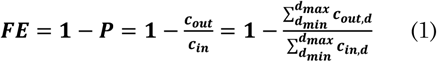

Where *P* is the penetration of particles through the filter, *c*_*in*_ refers to the particle concentration in the air entering the filter, *c*_*out*_ is the particle concentration exiting the filter, and *d* is particle diameter.

Figure 1 shows an overview of the measured filtration efficiencies of masks in this study, supplemented with results reported in other studies [30–34]. Here, no mask leakage was accounted for. It can be seen that filtration efficiency depends not only on the aerosol diameter but also highly varies for different masks and experimental tests. The fractional filtration efficiency typically drops around 0.1-0.5 µm. For this particle size, the filter collects the least number of particles [35].

**Figure 1.**
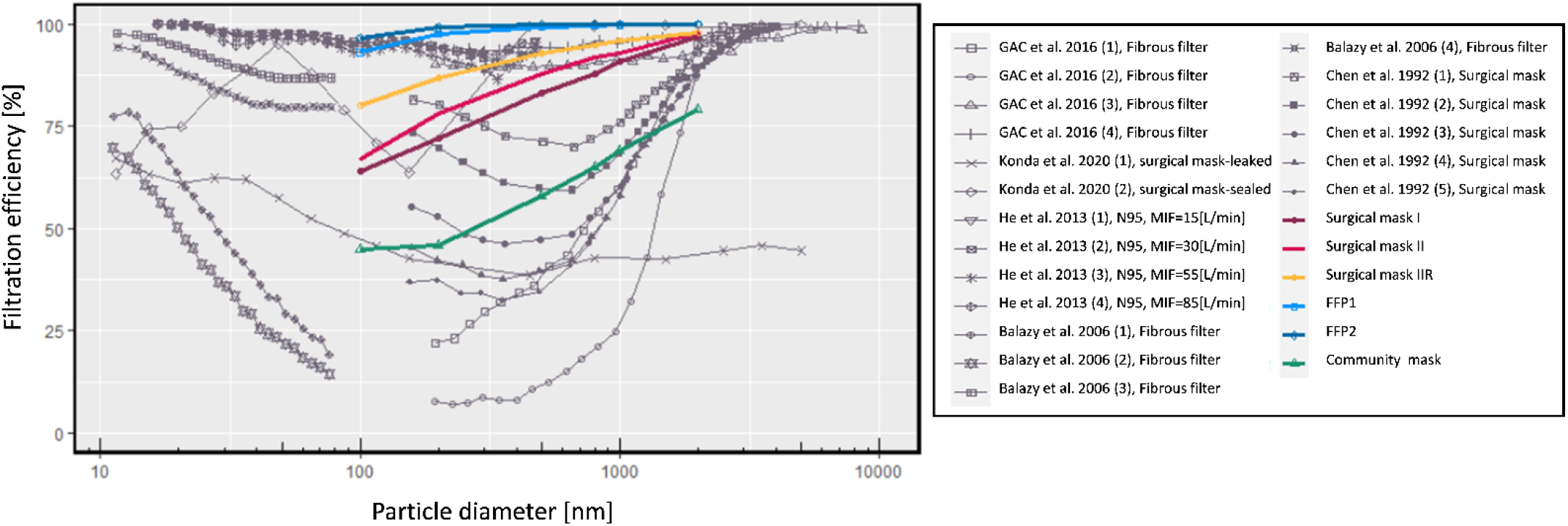
Measured filtration efficiency at different particle sizes [30–34]. The colored lines represent the filtration efficiencies of different types of masks that are being implemented in this study.

### 2.3 Masks and Standards

The mask characteristics and standards used to test masks are shown in Table 1.

**Table 1.**
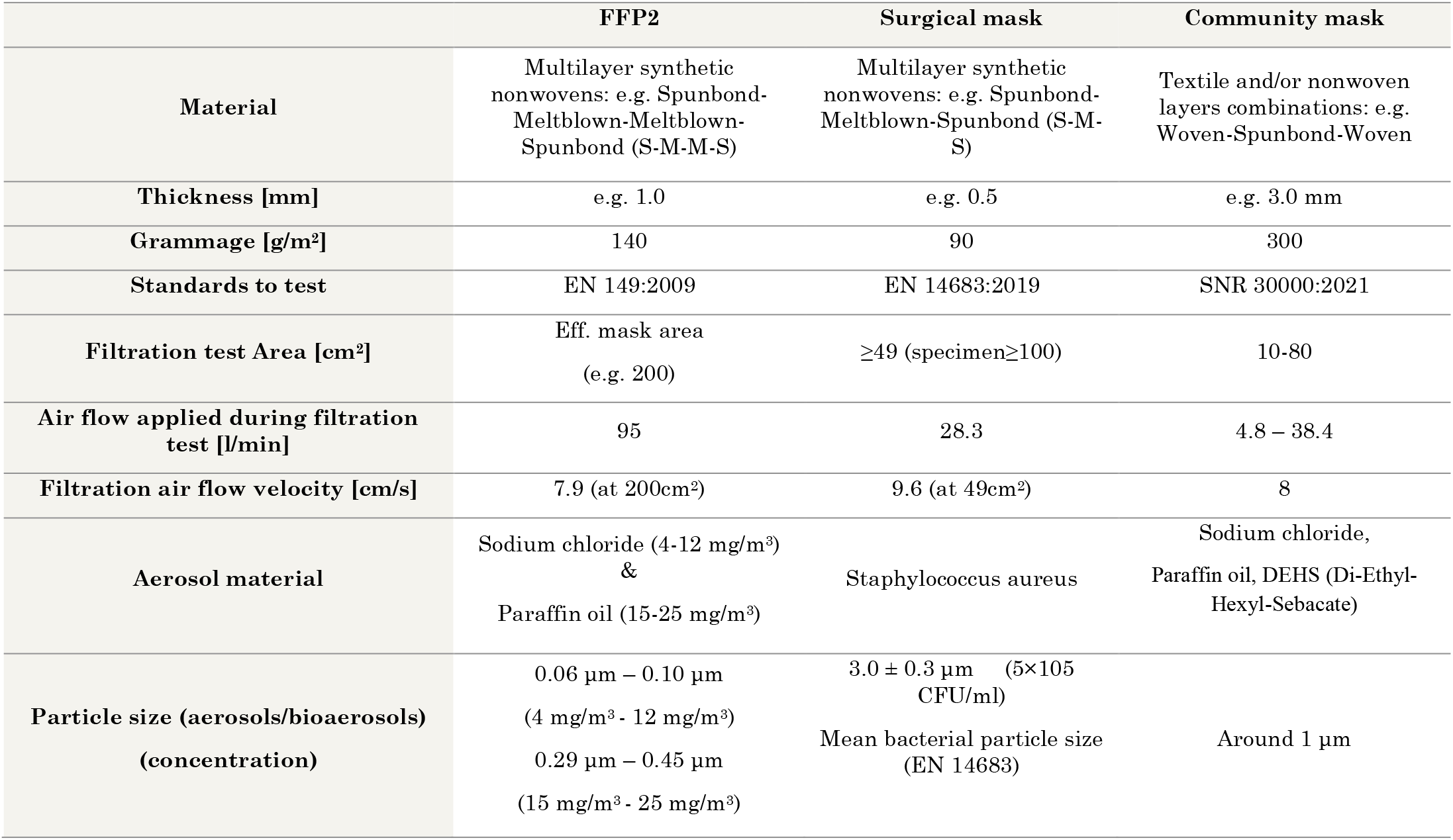
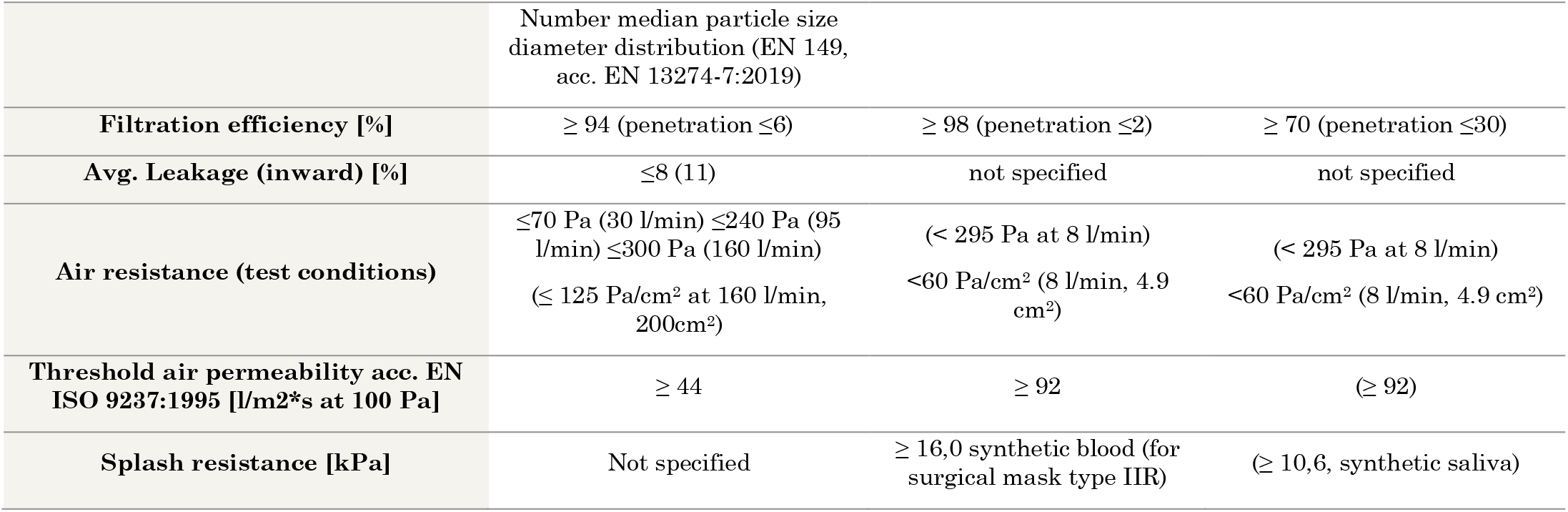
standards and characteristics of FFP2, Surgical mask, and Community mask.

## 3 Material and Methods

We mimicked different situations in which infected and healthy people are in close contact. To this end, we considered different breathing patterns of the people based on their activity level and whether or not they use a facemask. The environment in which they reside is also considered and characterized by the volume of the indoor environment and possible fresh air ventilation. In addition, the change in exposure is investigated for an infected person talking or coughing, which affects the number of emitted aerosols as well as the size distribution. We quantified the time needed to inhale a number of viruses to have a 50% chance of infection in a healthy person in a particular environment by considering all these factors. In Figure 2, the system’s overall structure considering these multiple factors is shown schematically.

**Figure 2.**
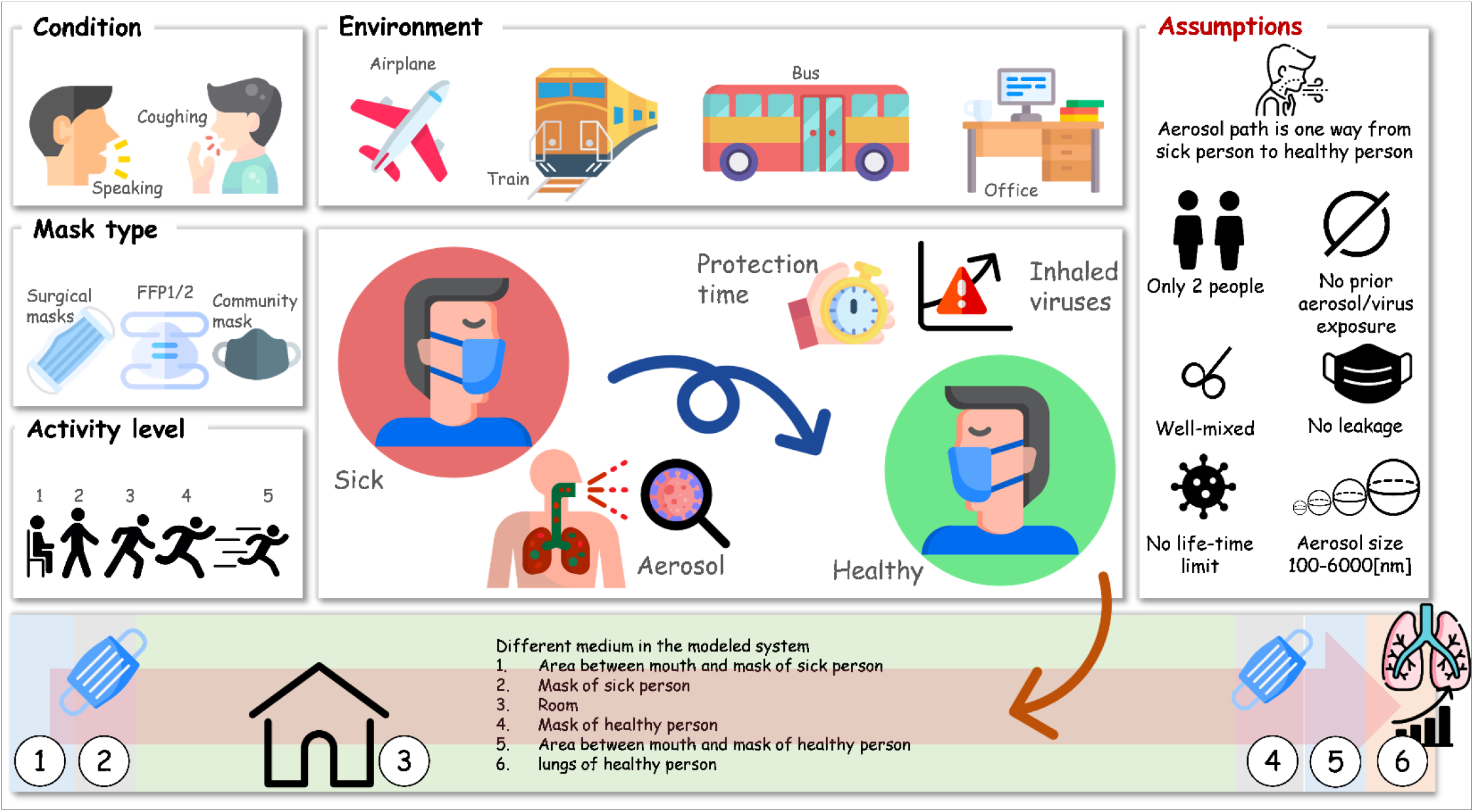
The overall structure of the modeling system, including breathing condition, mask type, environment, activity level, and modeling assumptions, is depicted here. (the icons are from https://www.flaticon.com/)

### 3.1 Experimental measurements of masks

#### 3.1.1 Mask air permeability experiment

Air permeability is one of the major factors in respect to the comfort of a facemask. High humidity levels inside the mask, which can accumulate over time of wearing, can lead to discomfort, which could result in the incorrect use of the mask. For facemasks, we assumed the airflow through the mask is laminar. This implies a linear relationship between the air flow rate through the mask (G_a_ [m^3^/s]) and the pressure drop (ΔP_i_ [Pa]). Therefore we can apply Darcy flow, with the permeability K [kg/(m^4^ s)] to the airflow (shown in Equation 2)

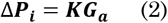

Since fit to the face of single-use masks is mostly not perfect, there is always a certain amount of air by-passing the filter material through leakage across the interface. Even leakage in the micrometer scale can be of relevance and become predominant in filter applications [36]. However, if leakage is reduced to a minimum, breathing resistance and humidity transport of the filter material become important factors for the comfort and safety of the mask. It should be noted that in this study, the airflow from the opening parts between the face and facemask is not considered.

The pressure drop (breathing resistance) of an ideally homogenous fiber filter material is shown in Equation 3.

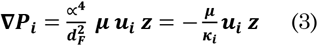

In which α is packing density (fiber material density / flat sheet density), z is the characteristic length (filter media thickness), d_F_ is the circular fiber cross-section (fineness) [m], u_i_ is the air velocity [m/s]], κ_i_ is the air permeability through the mask [m^2^], and μ is the gas viscosity [Pa.s]. As the breathing resistance of the mask (α^4^ µ z /d^F2^) decreases, the breathability of the mask increases. Consequently, a critical aspect of good breathability of mask materials is to minimize packing density while maintaining filtration efficiency. The best way to achieve this is to use fine fibers at a relevant thickness simultaneously. The diameter of fibers in common facemasks is between 0.5-10 µm [37]. Airflow resistance for a continuous airflow can be measured in calibrated devices described in EN 14683 or with air permeability test benches (ISO 9237). If air permeability test devices are used, it is recommended to use the sample size requested from the mask test standard and to re-calculate the pressure over the given test surface area from the set test pressure. The pressure resistance for the different types of masks included in this study is provided in Table 2.

**Table 2.** pressure resistance for surgical, FFP, and community masks based on EN 14683 standards.

| Mask type | Surgical mask (I) | Surgical mask (II) | Surgical mask (IIR) | FFP1 | FFP2 | Community mask |
| --- | --- | --- | --- | --- | --- | --- |
| Standard | EN 14683 |  |  |  |  |  |
| Conditions | Surface area: 4.9cm<sup>2</sup>, Temperature: 21°C, Relative humidity: 85%, Airflow: 8 L/min |  |  |  |  |  |
| Pressure resistance [Pa/cm<sup>2</sup>] | 40 | 41 | 39 | 54 | 84 | 56 |

#### 3.1.2 Mask filtration efficiency experiment

To perform filtration efficiency tests, a circular specimen with a diameter of 60 mm was sampled from a mask and sealed airtight in a sample holder to obtain an effective test surface of 1.66*10^3^ mm^2^. An aerosol was drawn with an aerosol generator (AGK2000) from a solution of 0.02 g mL^-1^ of fructose in demineralized water. Fructose particles at a concentration of 35 mg m^-3^ in dried air were neutralized in a corona discharging unit (CD2000) and driven to the sample. A constant airflow of 8 L min^-1^ was set through the mask specimen (from outside to inside). Particle penetration through the sample was quantified using the particle analyzer Combustion DMS500). This ‘Fast Particulate Spectrometer’ uses unipolar corona charging and parallel detection of particles of varying electrical mobility to offer real-time measurement of the particle size spectrum between 5 and 2,500 nm. The filtration efficiency was determined by comparing a steady flow of particles after a constant concentration was reached for 2 minutes with the mask sample and afterward measuring the raw gas concentration for another minute without the mask sample. This filtration efficiency (FE) was expressed as a percentage and was reported in the particle range from 100 nm to 2000 nm, based on triplicate measures. The filtration efficiency for specific aerosol diameters for the different masks included in this study is presented in Table 3. The community mask in this study is 100% polyester, which is equipped with an anti-bacterial treatment. This mask has a splash resistance at pressure of 12 kPa based on ISO 22609 and air pressure resistance of <70 Pa cm^-2^ based on EN 14683.

**Table 3.** Filtration efficiency of different types of masks

|  | Surgical mask (I) | Surgical mask (II) | Surgical mask (IIR) | FFP1 | FFP2 (N95) | Community mask |
| --- | --- | --- | --- | --- | --- | --- |
| Particle size [nm] | Filtration efficiency [%] |  |  |  |  |  |
| 100 | 64 | 67 | 80 | 93.1 | 96.6 | 45 |
| 200 | 72 | 78 | 87 | 97.5 | 99.1 | 46 |
| 500 | 83 | 88 | 93 | 99.2 | 99.8 | 58 |
| 800 | 88 | 92 | 95 | 99.6 | 99.9 | 65 |
| 1000 | 91 | 93 | 96 | 99.8 | 99.9 | 69 |
| 2000 | 97 | 98 | 98 | 99.9 | 99.9 | 79 |

### 3.2 Physics-based model for virus filtration

#### 3.2.1 Computational system configuration

This study modeled a rectangular one-dimensional domain that represents a fraction of a mask with a length of 2.5 [cm], similar to the standard experiments, and a width of 0.5[mm]. The air permeability coefficients for mask types were calculated based on experimental data (Table 2). With this, the velocity of the air and the total air exhaled/inhaled air volume was calculated dependent on the breathing rate to get the total number of aerosols. As air goes solely through the mask, the mask will filter a fraction of these aerosols based on their diameter. As such, only a part of these particles will penetrate through the mask and reach the surrounding area. This study assumed that any aerosol passing the mask would not be inhaled again by the infected person but would remain in the environment. The environment was considered to be instantly well-mixed, implying that the concentration of particles in the environment is uniform at all times. On the other hand, a healthy person will inhale the accumulated aerosols in the environment. If this healthy person is wearing the mask, only a fraction of these aerosols will reach the respiratory tract. To calculate the number of aerosols that are transferred from infected to healthy persons, we considered six instantly well-mixed domains in the overall modeling environment: 1. The zone between the respiratory airway and mask for the infected person, 2. The mask characteristics of the infected person, 3. the characteristics of the room in which these two people are meeting, 4. The mask characteristics of the healthy person 5. The zone between the respiratory airway and mask for the infected person, and 6. The inner airway of a healthy person (Figure 3).

**Figure 3.**
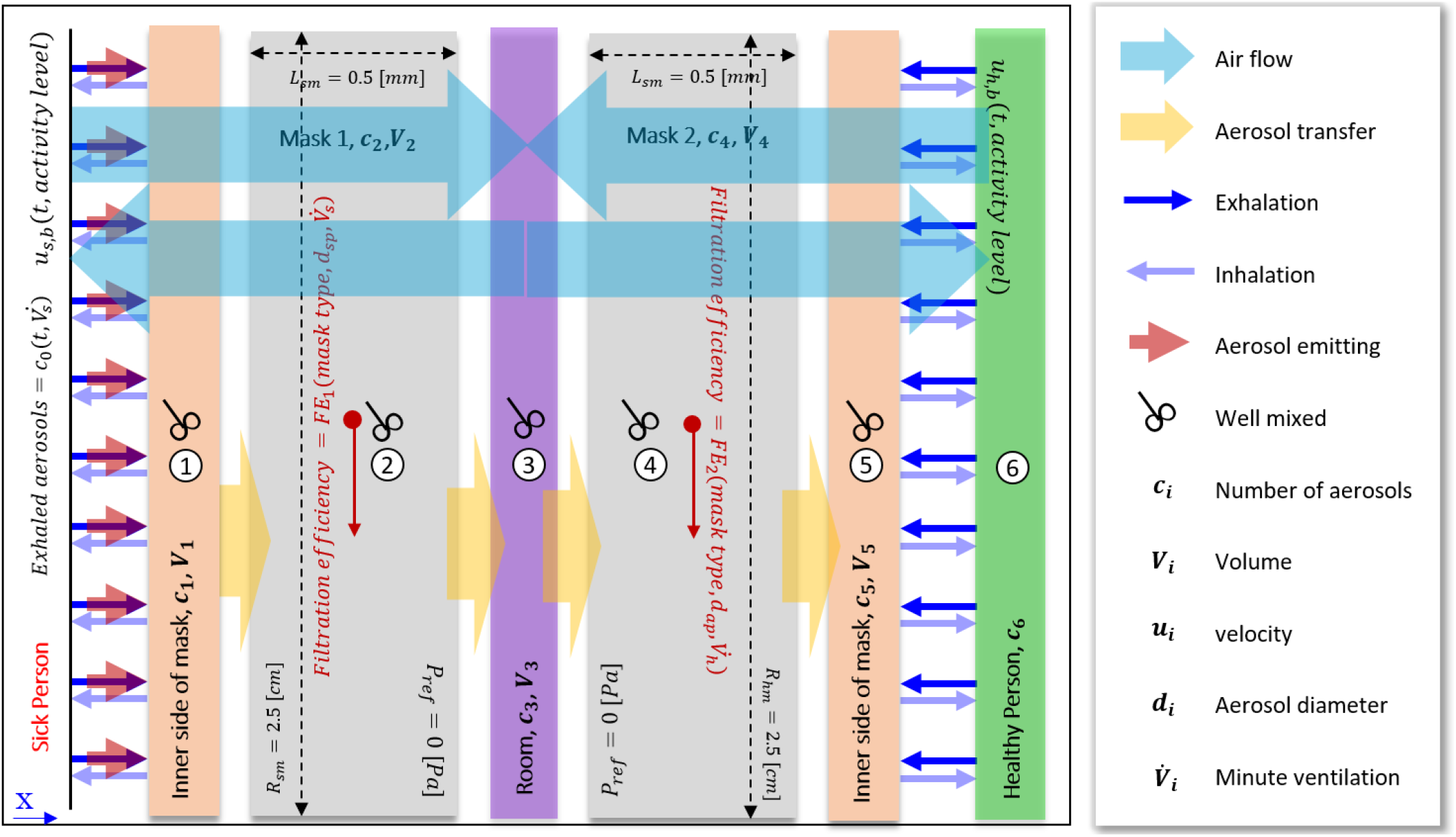
Geometry and boundary conditions of the overall model for two people (infected and healthy people) in close contact. The modeled geometry consists of 6 zones: the zone between the respiratory airway and mask for the infected person; the mask of the infected person; the room in which these two people are meeting: the mask of the healthy person; the zone between the respiratory airway and mask for the infected person; and the inner airway of a healthy person

#### 3.2.2 Governing equations

##### Airflow

The experimental data shows a linear relationship between airflow through the filter of the mask and the pressure drop. This relation implies that Darcy’s law can predict the airflow behavior through the mask in the operational range of airspeed for breathing which is brought in Equation 3. The continuity is applicable for airflow, and it is given by Equation 4.

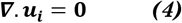

##### Aerosol transport

The contaminated particles emitted by an infected person during breathing, coughing, and speaking will be partially filtered by the facemask, whereas the remaining will be spread into the environment, where healthy people can inhale them. To evaluate the fraction of filtrated aerosols in the mask, we considered filtration efficiency as a function of mask type and aerosol diameter. The mass conservation for the number of aerosols is mentioned in the following equations (5-10). As mentioned earlier, we considered six different zones, for which we solved each transport equation for the particles (Figure 3): 1. The inner side of the mask for the infected person (Eq. 5), 2. mask of the infected person (Eq. 6), 3. Room (Eq. 7), 4. Mask of the healthy person (Eq. 8), 5. The inner side of the mask for the healthy person (Eq. 9), and 6. Healthy person’s airway (Eq. 10). At the boundaries, we assume continuity of the particle fluxes.

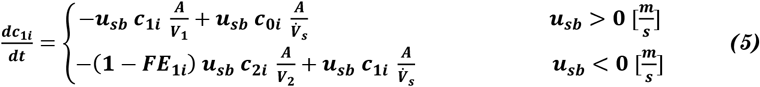

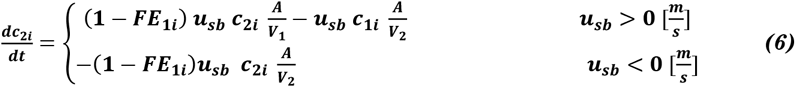

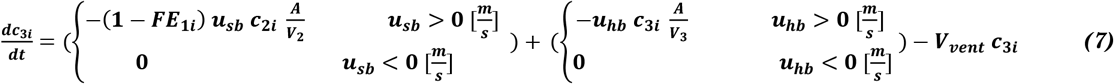

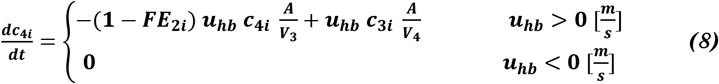

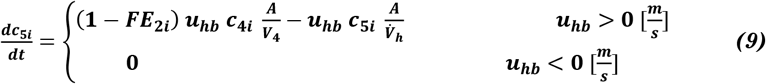

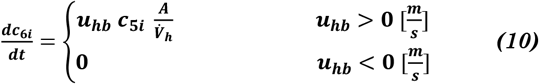

Where c_i_ is the number of aerosols in medium i (shown in Figure 3), u_sb_ and u_hb_ are the air velocity of breathing for the infected person and healthy person, respectively. The surface area is shown by A, and V_i_ is the volume of medium i. In these equations, FE_1i_ refers to the filtration efficiency of the infected person’s mask, and FE_2i_ refers to the filtration efficiency of the healthy person’s mask.

##### Viral load per aerosol

A variety of respiratory viruses like influenza virus and acute respiratory syndrome (SARS) coronavirus are available in high concentrations in infected human saliva and respiratory mucus. The produced aerosols from human saliva will carry these viruses by breathing, talking, and coughing. The virus content of these aerosols is dependent on the aerosol’s diameter. Based on the Zuo et al. study, the number of MS2 bacteriophages [plaque-forming unit (PFU)] per aerosol based on its diameter is shown in Equation 11.

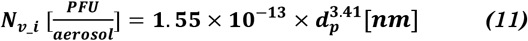

Where N_v_i_ are the number of bacteriophages per aerosol and d_p_ is the aerosol’s diameter [38]. As MS2 bacteriophages are nonpathogenic, it is one of the most common human-pathogenic virus surrogates and they are used in various viral studies [38]. Due to the lack of similar data for SARS-COV-2, we assume that the condition for the viral load of SARS-CoV-2 in the exhaled aerosols by an infected person is similar to MS2 bacteriophages (Equation 11).

#### 3.2.3 Material properties and transport characteristics of mask and environment

#### 3.2.4 Boundary and initial conditions

On the inner side of the mask for the infected and healthy person, the airflow at the boundary condition is time and activity level dependent. The airflow intensity and pattern based on time and activity level are shown in Table 4. The exhaled breath of the infected person is the source of aerosols, which depends on the volume of exhaled air that enters the room by expiration [39]. The initial number of virus-laden aerosols in the entire system (the six blocks shown in Figure 3) is set to zero.

**Table 4.**
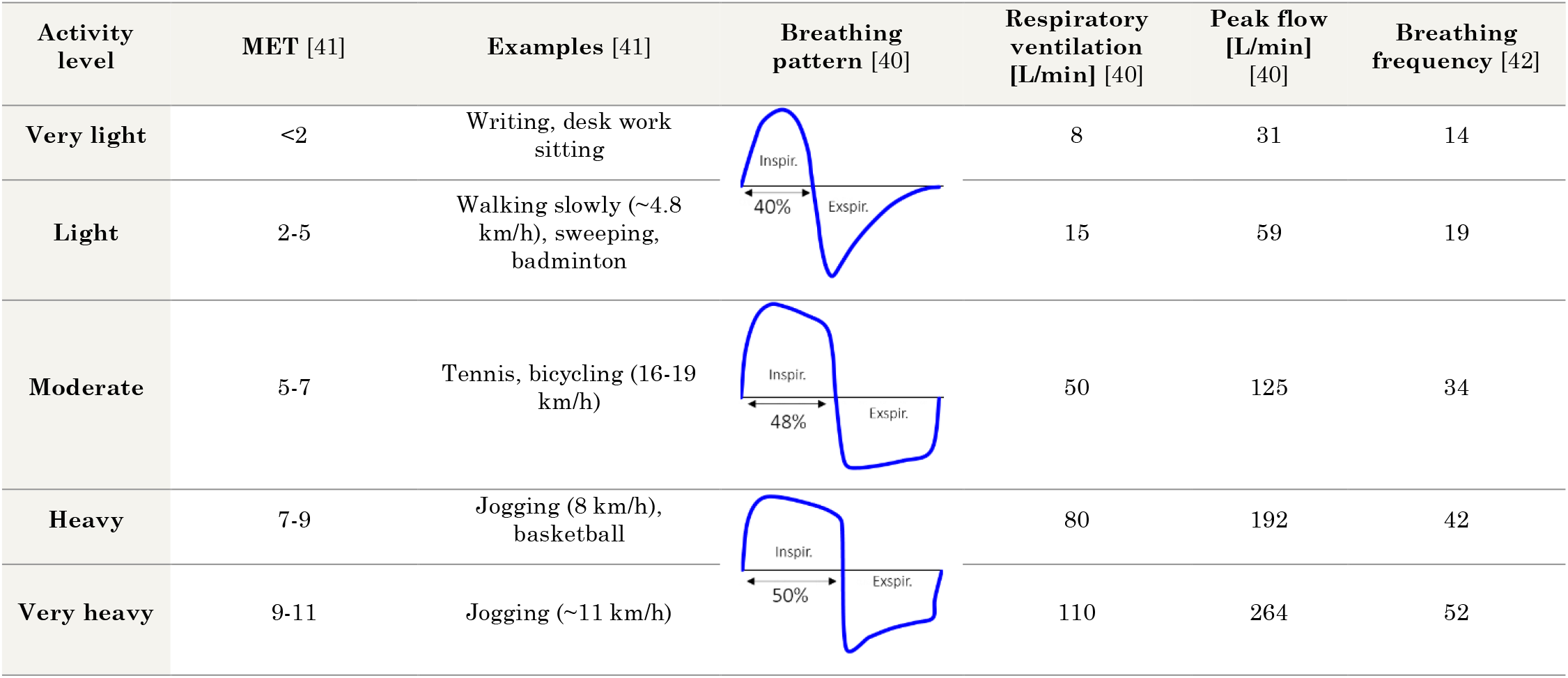
Breathing pattern based on activity level.

| Activity level | MET [41] | Examples [41] | Breathing pattern [40] | Respiratory ventilation [L/min] [40] | Peak flow [L/min] [40] | Breathing frequency [42] |
| --- | --- | --- | --- | --- | --- | --- |
| Very light | <2 | Writing, desk work sitting |  | 8 | 31 | 14 |
| Light | 2-5 | Walking slowly (~4.8 km/h), sweeping, badminton |  | 15 | 59 | 19 |
| Moderate | 5-7 | Tennis, bicycling (16-19 km/h) |  | 50 | 125 | 34 |
| Heavy | 7-9 | Jogging (8 km/h), basketball |  | 80 | 192 | 42 |
| Very heavy | 9-11 | Jogging (~11 km/h) |  | 110 | 264 | 52 |

### 3.3 Simulated configurations

Human exposure is simulated in different environments such as an office room, a bus, a train, and an airplane. The properties of these rooms are detailed in Table 5. The volume of each room is modified based on the capacity of the room to find the effective volume for the situation when the infected person is sitting next to a healthy person. In such a way, if the bus cabin volume is roughly 109 m^3^, and the capacity of the bus is 77 people, by dividing the volume by every two people, we reach the number of 2.5 m^3^, which is reported in Table 5. In this regard, the volume of the train, bus, and airplane cabin is divided by their capacity to calculate the volume when these two people are in close contact as a worst-case scenario.

**Table 5.** properties of the modeled rooms

| Room | Office room | Train | Bus | Airplane |
| --- | --- | --- | --- | --- |
| Volume [m<sup>3</sup>] | 28 | 150 | 109 | 163 |
| Capacity | 2 | 80 | 77 | 110 |
| Volume* [m<sup>3</sup>]<br>(normalized for 2 people) | 28 | 4 | 2.5 | 3 |
| Model | - | SBB RABe 511 | Neoplan Centroliner N4516 standard bus | A220-100 |
| Ventilation rate [1/h] | 7 | 11 | 4 | 25 |
| Ref. (for air ventilation rate) | [43] | [44] | [45] | [46] |
| * The volume of each environment modified based on the capacity |  |  |  |  |

In this study, we assumed additional varieties such as talking and coughing. When the person starts to talk or cough, the emitted aerosol’s total number and size distribution will change. However, this can vary between individuals, the intensity, or even what sort of words are expressed. To reduce the complexity, we decided to only consider an average value for the total number of emitted aerosols during talking and coughing. Based on previous studies, for particles in the range of 500 [nm] to 20 [µm], the total number of emitted aerosol during talking and coughing is three times and 20 times higher than regular breathing, respectively [47,48]. A repetitive speaking-breathing pattern of 10 [s] talking followed by 10 [s] of breathing was applied. For coughing, the data was extracted from the Johnson et al. study in which volunteers had a mild throat-clearing cough intensity with a continuous frequency while they were comfortable during 30 [s][21]. We additionally considered three more scenarios for coughing, which imply the situations when the infected person coughs 50%, 25%, or 10% of the mentioned frequency for coughing.

### 3.4 Numerical implementation and simulation

COMSOL Multiphysics® software (version 5.6, COMSOL AB, Stockholm, Sweden), a finite-element-based commercialized software, was used in this study. The airflow simulation was done using equation-based modeling to solve Darcy’s law and continuity, modeled by the ODE interface in COMSOL. The particle transfer was modeled by assuming well-mixed domains and was modeled by using the ODE interface in COMSOL. Quadratic Lagrange elements were used with a fully coupled direct solver, which relied on the MUltifrontal Massively Parallel sparse direct Solver (MUMPS) solver scheme. In order to capture the fluctuation of airflow, aerosol and viral density of the environment, the time step of 0.01 [min] was chosen. The tolerances for solver settings and convergence were determined by means of sensitivity analysis in such a way that a further increase in the tolerance did not alter the resulting solution.

### 3.5 Metrics

Based on the minute breathing rate 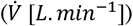 the total volume of ventilated air at each time step for each person is calculated, as in Equation 12.

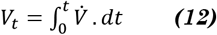

Which, *V*_*t*_ is the total volume of ventilated air up to time t. The total number of emitted and inhaled aerosols and corresponding virus particles are calculated with Equations 13-14.

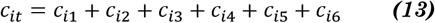

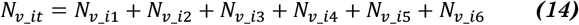

The protection time in this study is defined as the time when the number of inhaled viruses in a healthy person reaches a particular threshold. This threshold is defined by ID_50_ representing the minimum number of viruses needed to infect 50% of persons exposed. Due to the lack of sufficient data for SARS-CoV-2, we assumed the ID_50_ of Influenza A (790 [49]) is the same as SARS-CoV-2. Besides the number of inhaled particles, the time window of exposure can also affect the possible infection. Given the limited information on this effect on the infection risk, we did not consider this.

## 4 Results and Discussion

### 4.1 Air ventilation

As mentioned in section 3.3, regular air ventilation of the room is considered to limit the increase (Equation 7) in the concentration of the aerosols in the room. To investigate the effect of ventilation on the total number of aerosols in the environment, the office room (detail in Table 5) was studied by considering five different activity levels in 2 conditions with and without air ventilation. The result is shown in Figure 4. Based on the result, when we have air ventilation in the office room, by increasing the activity level from 1 to 5, in 30 minutes, the total number of aerosols increases by about 2600 times. On the other hand, if the office room does not have any air ventilation, the total number of aerosols in the environment during 30 minutes for different activity levels will increase to about 100 times more compared to the room with air ventilation. Therefore, the lack of air ventilation can considerably increase the risk of infection even considering the lowest activity level.

**Figure 4.**
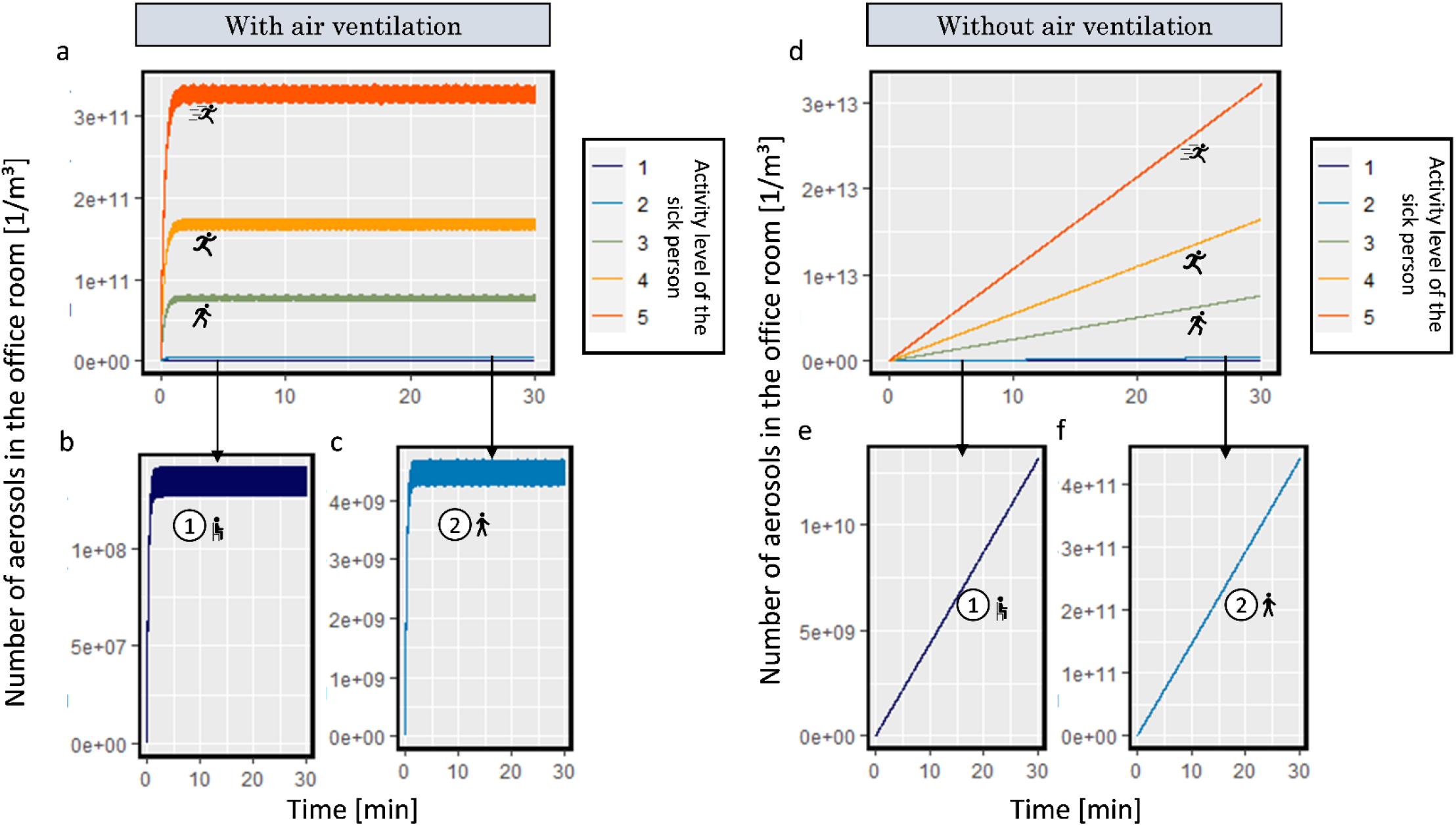
a. Number of emitted aerosol in the office room with air ventilation during 30 minutes by 5 different activity levels; b. activity level 1; c. activity level 2; d. Number of emitted aerosol in the office room without air ventilation during 30 minutes by five different activity levels; e. activity level 1; f. activity level 2. (the icons are from https://www.flaticon.com/)

### 4.2 Contamination of the environment with virus-laden particles by an infected person for different activity levels and mask types

This section quantifies how much infected-person contaminates their environment and how much facemasks can help avoid this contamination. A SARS-CoV-2 infected person constantly emits respiratory aerosols during breathing and also droplets during talking or coughing, all of which contain viruses. These exhausted virus-laden aerosols spread the virus and put healthy people at risk. The diameter distribution of aerosols we have used in this study is based on the literature shown in Figure 5a. By considering different activity levels, we map the total emitted number of aerosols and viruses from an infected person for 30 [min]. As the person’s activity level increases, the exhaled air volume increases too. By increasing the volume of air, the number of exhausted aerosols increases. Figure 5b,c shows the total volume and the total number of exhaled aerosols based on the activity level. The total number of exhaled aerosols for activity level 1 (such as sitting) during breathing after 30 [min] is equal to 1.28*10^12^. Each aerosol can carry a different number of viruses based on its size. Based on the aerosol size distribution and the relationship between the aerosol’s initial volume and viral load, the total number of emitted viruses is shown in Figure 5d. The effect of facemasks in reducing the total number of emitted aerosols to the environment for different activity levels compared to not wearing a mask in blocking aerosols is shown in Figure 5/e, g, i, k, and m. The corresponding effect on the emission of viruses is shown in Figure 5/j, f, h, j, l, and n.

**Figure 5.**
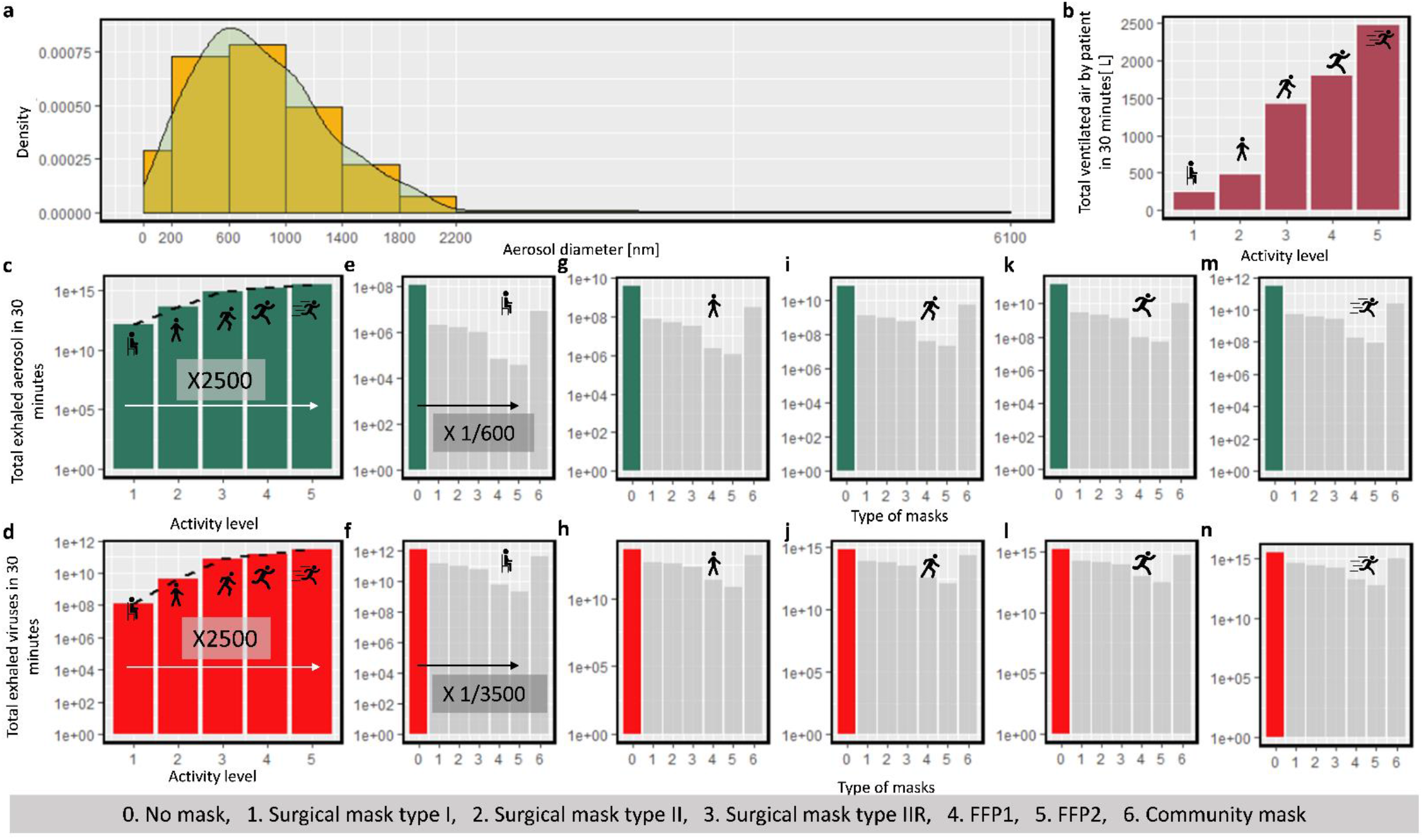
a. diameter distribution of exhaled aerosols by the infected person, b. accumulated ventilated air by the infected person, c, e, g, i, k, and m. total exhaled aerosols d, f, h, j, l, and n. total exhaled viruses during five activity levels (sitting, walking, moderate activities, running, and sprinting) by using no mask (0), surgical mask I (1), surgical mask II (2), surgical mask IIR (3), FFP1 (4), FFP2 (5), and community mask (6) by the infected person in 30 [min]. (the icons are from https://www.flaticon.com/)

The number of emitted aerosols to the environment increases with more strenuous activity levels and corresponding increased breathing activity. As the total number of emitted aerosols for activity level 1 (only breathing) during 30 [min] is 1.28*10^12^ while this number is 3.14*10^15^ (almost 2500 times more) for high exertions. This implies that the high activity of the infected person increases the virus concentration in the environment. During this 30 [min], 99.8% of the total number of aerosols and 99.9% of the number of viruses were blocked by the FFP2 mask. For the same scenario, surgical mask type I, which has the lowest filtration efficiency among the standardized masks, blocked 89.4% of aerosols and 95.9% of viruses. The filtration efficiency of FFP2 is 11.6% more than surgical mask type I, while FFP2 viral blockage is only 4.2% more than surgical mask type I. On the other hand, the FFP2 mask reduced the number of emitted aerosols by 600 times, while it reduced the number of viral particles by 3500 times. By comparison of these numbers, we realize that the mask must have a high filtration efficiency for aerosols that have a higher frequency (200-1000 [nm]) and aerosols which carry a higher number of viral copies (>2000 [nm], as they are larger, they can carry more viruses). Therefore, the overall average filtration efficiency of a mask does not give us how effective this mask is for reducing the viral contamination of the environment.

### 4.3 Protection time of different mask types for a healthy person

A specific number of viruses need to enter the body via the respiratory tract of each individual to infect that person. In a healthy population, the risk probability of infecting 50% of people is defined by the ID_50_ value. As different aspects of SARS-CoV-2, including ID_50,_ are still unknown, we used the reported ID_50_ for the Influenza A. virus and analyzed different conditions affecting reaching this threshold.

#### 4.3.1 Effect of breathing condition on mask protection time

Different conditions, such as speaking and coughing, change the distribution and number of emitted aerosols, which is shown in Figure 6/a. The protection time while an infected person might start to talk or cough is shown in Figure 6/b. We considered four different scenarios in an office room: 1. None of them wear masks 2 & 3. one of them wears surgical masks IIR, and 4. Both of them wear surgical masks IIR. The maximum protection time we considered in this study was to be 8 [h] (referring to a maximum residence time in the room), and we did not continue analyzing the performance after this time. Based on the result, if the infected and healthy persons both wear a mask, the protection time at each condition is over 8[h]. We did not observe any differences in scenarios 2 and 3, where only one person wore the surgical mask. This similarity can be explained by the one-way transmission of the virus, which we assumed in this simulation. If the infected person constantly coughs, the protection time reduces to less than 20% of the time the infected person does not cough and speak. Even with a lower frequency of coughing (Coughing-4, which has 10% of the frequency of the maximum number of coughs in a defined time), the protection time is reduced by 40%, which is similar to the reduction of protection during talking in the condition without a mask.

**Figure 6.**
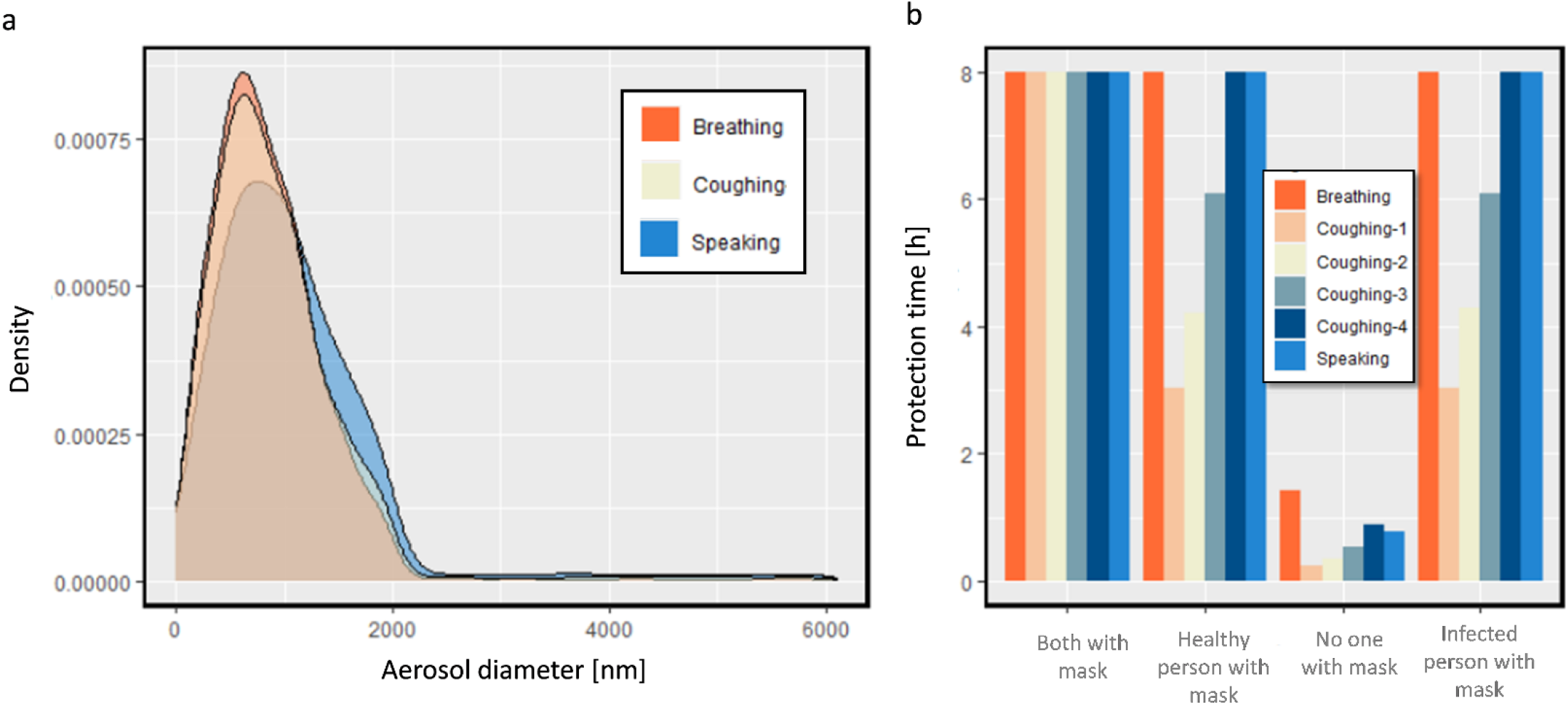
a. Diameter distribution of exhaled aerosols by the infected person during breathing, speaking, and coughing, b. protection time for a healthy person in an office room.

#### 4.3.2 Effect of activity level on mask protection time

We quantified how variations in the breathing pattern by changes in the activity level affect the number of emitted aerosols (section 4.1) and the protection time of different masks. In this regard, we considered the same four scenarios: 1. none of the people (infected and healthy people) wears a mask, 2. only the infected person wears a surgical mask IIR, 3. only the healthy person wears a surgical mask IIR, and 4. both of them wear surgical mask IIR. The environment for this study is a standard office room with a modified volume of 28 m^3^. As is shown in Figure 7, when both people increase their activity level from 1 to 2 (e.g., from sitting to walking), the protection time decreases by more than 90% when only one of them wears the mask. Suppose they increase their activity to higher levels; in that case, the protection time decreases drastically for the maximum activity; even if both of them wear the surgical mask IIR, the protection time remains only to be 10 [min]. It should be noted that when the protection time is so low (lower than 10 min), the condition does not meet the requirement for a well-mixed assumption. Therefore, the evaluated protection time for these conditions might considerably deviate from reality. In this case, the protection time depends on the distance and location of healthy and infected persons can be higher or lower. However, it still can be concluded that the protection time when both people have high activity is considerably low. This can be an issue in places where people might engage in demanding activities, like gym and indoor work with high physical demand.

**Figure 7.**
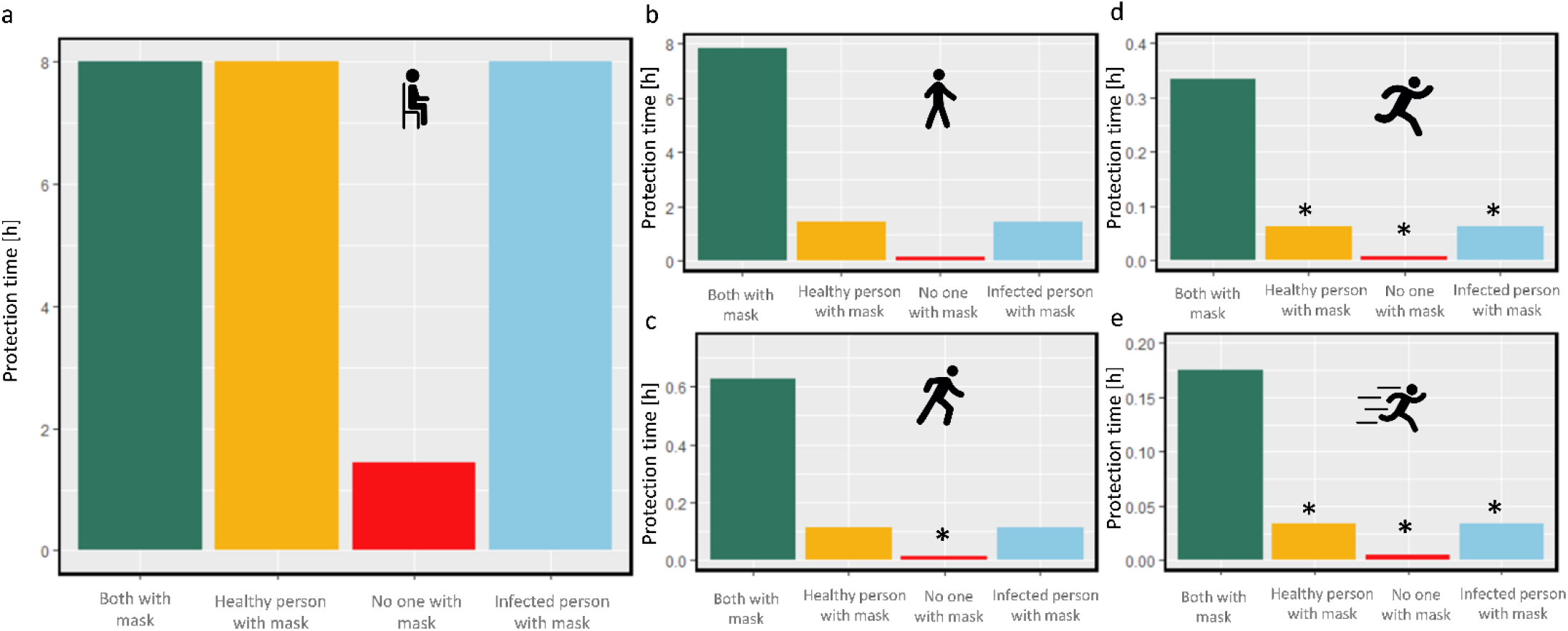
protection time for a healthy person in an office room during different activity levels. a. level 1 (e.g., sitting), b. level 2 (e.g., walking), c. level 3 (e.g., climbing), d. level 4 (e.g., running), e. level 5 (e.g., sprinting), with different conditions with and without the mask. The star represents the too low protection time which is not compatible with the well-mixed assumption. (the icons are from https://www.flaticon.com/)

#### 4.3.3 Effect of room enclosure on mask protection time

Contact between a healthy and infected person can occur in different environments such as office rooms, buses, trains, and airplanes. As the aerosols get emitted by an infected person to the environment by considering an instantaneously well-mixed medium, their concentration gets diluted by the volume of the environment. If the environment has air ventilation, the dilution is further increased. Based on the studied environment, We only considered low activities such as sitting, while the infected person might speak or cough at different frequencies. In this section, a surgical mask IIR was implemented. Three different scenarios of wearing the mask were studied: 1. None of them is wearing the mask, 2. One of them is wearing a mask, and 3. Both of them are wearing masks. In Figure 8, the result is shown by considering the average residence time for each environment (horizontal green line). Based on the result, if both people do not wear a mask, the protection time is shorter than the average residence time in all the environments. If only one of them wears the mask while breathing, the healthy person is protected longer than the average staying time. However, when the infected person starts to cough, the protection time drops, which might put the healthy person in danger. In every case, the healthy person can have a lower chance of getting infected in these environments for the assumed time if both of them wear the mask. Based on the result, in the bus by only breathing, when only one of them wears the surgical mask IIR the protection time is 1.3 h, while if both of them wear a facemask, the safe exposure duration increases by 4.5 folds to 7.1 h. If both people wear a facemask on the train by only breathing, the protection time is 8 h, while if only one wears it, this number is only 2.8 h. On an airplane, by only breathing, the protection time for both people wearing masks is 8 h, and if only one of them wears a facemask is 2.1[h]. It should be considered that as the infected person starts to talk or cough, the risk for the healthy person increases considerably. Therefore, it is better if both people wear a facemask to reduce the risk of infection in conditions such as coughing, speaking of the infected person, or staying on longer journeys.

**Figure 8.**
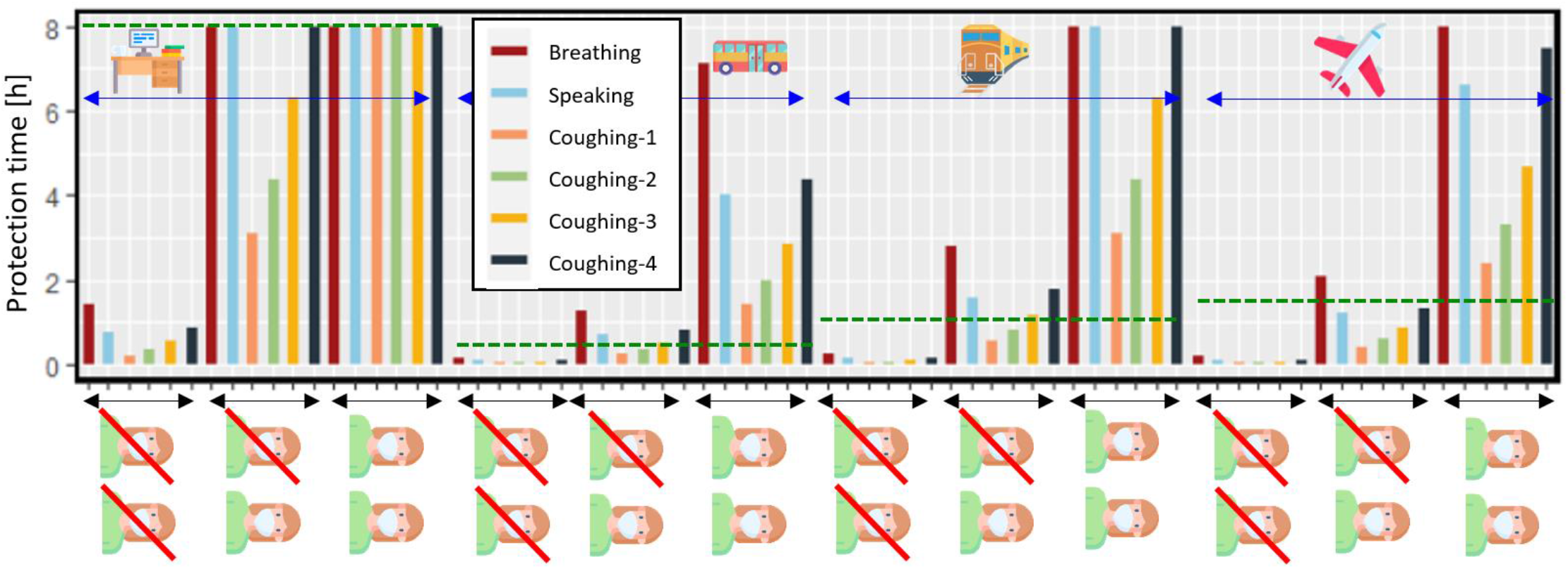
protection time for a healthy person in close contact with an infected person in office room, bus, train, and airplane, with or without surgical mask IIR, by considering the infected person’s breathing, coughing, and speaking. The dashed green line represents the average staying time in each environment. (the icons are from https://www.flaticon.com/)

#### 4.3.4 Effect of leakage of the mask on reaching the threshold

So far, the entire simulation was executed by the assumption of a fully sealed mask. Based on this assumption, all the exhaled and inhaled air passes through the mask, which filters the aerosols. However, this deviates from reality, as poorly fitting masks and improper use of masks must be considered. This section considers the impact of leakage on the filtering performance of surgical mask IIR, FFP2, and Community masks. The filtration efficiency for these three masks by considering the sealed and unsealed full-size masks on Sheffield’s head is shown in Figure 9/a. There is a difference between the filtration efficiency of sealed masks in this section and the filtration efficiencies considered for the same type of masks in previous simulations. This variation is due to different batches of masks used for this investigation. This highlights the variability in filtration efficiency to be considered even within the same type of face mask. By analyzing the filtration efficiency of sealed and unsealed facemasks, a considerable drop in apparent filtering is observed for all types of masks. This drop was more drastic for surgical mask type IIR. However, the filtration efficiency of the mask is similar regardless of the sealing. As a result of the gap between the face and the mask, a part of air enters and reaches the room without being filtered through the mask. Therefore the apparent filtration efficiency of the unsealed facemask is lower than the sealed masks. The different levels of impact of leakage on the apparent filtration efficiencies of these masks could be related to their shapes and fitting to the face. In section 4.2., the result showed that the performance that FFP2 provides is considerably higher than that of the surgical mask type IIR. Furthermore, the performance of surgical mask IIR is considerably higher than the community mask. The result in Figure 9/b for an office room, while both people wear the same type of mask, shows that the performance of FFP2 is almost in the same range as for surgical mask IIR and community mask by considering the unsealed condition. Additionally, the protection time for an unsealed surgical mask is lower than for an unsealed community mask. This implies that besides the high filtration efficiency of the surgical mask IIR, it cannot protect the wearer as expected due to its poor face sealing.

**Figure 9.**
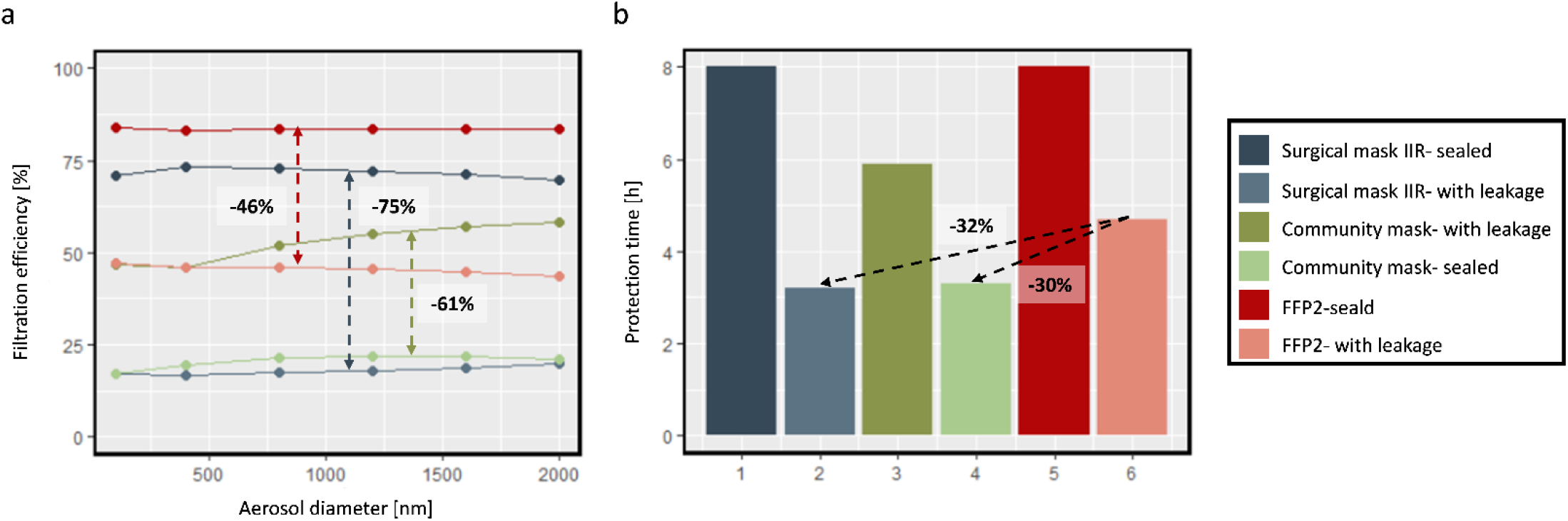
a. filtration efficiency for the surgical mask IIR, FFP2, and community mask by considering leakage, b. protection time while the healthy and infected people both wear the same mask in an office room with ventilation.

### 4.4 Virtual mask tester

We transformed our physics-based model of facemask into an openly accessible computer program named ‘the Virtual Mask Tester’ (Figure 10). This application is suitable for users interested in evaluating the performance of masks in different conditions. The mask can be the standard or newly developed masks for personal or production uses. The user can choose different environments for the study. Besides the defined environment in this study, they can define a room with custom volume and air ventilation. Users can even evaluate their own masks with a custom particle-size-dependent filtration efficiency. The link to access the application is here.

**Figure 10.**
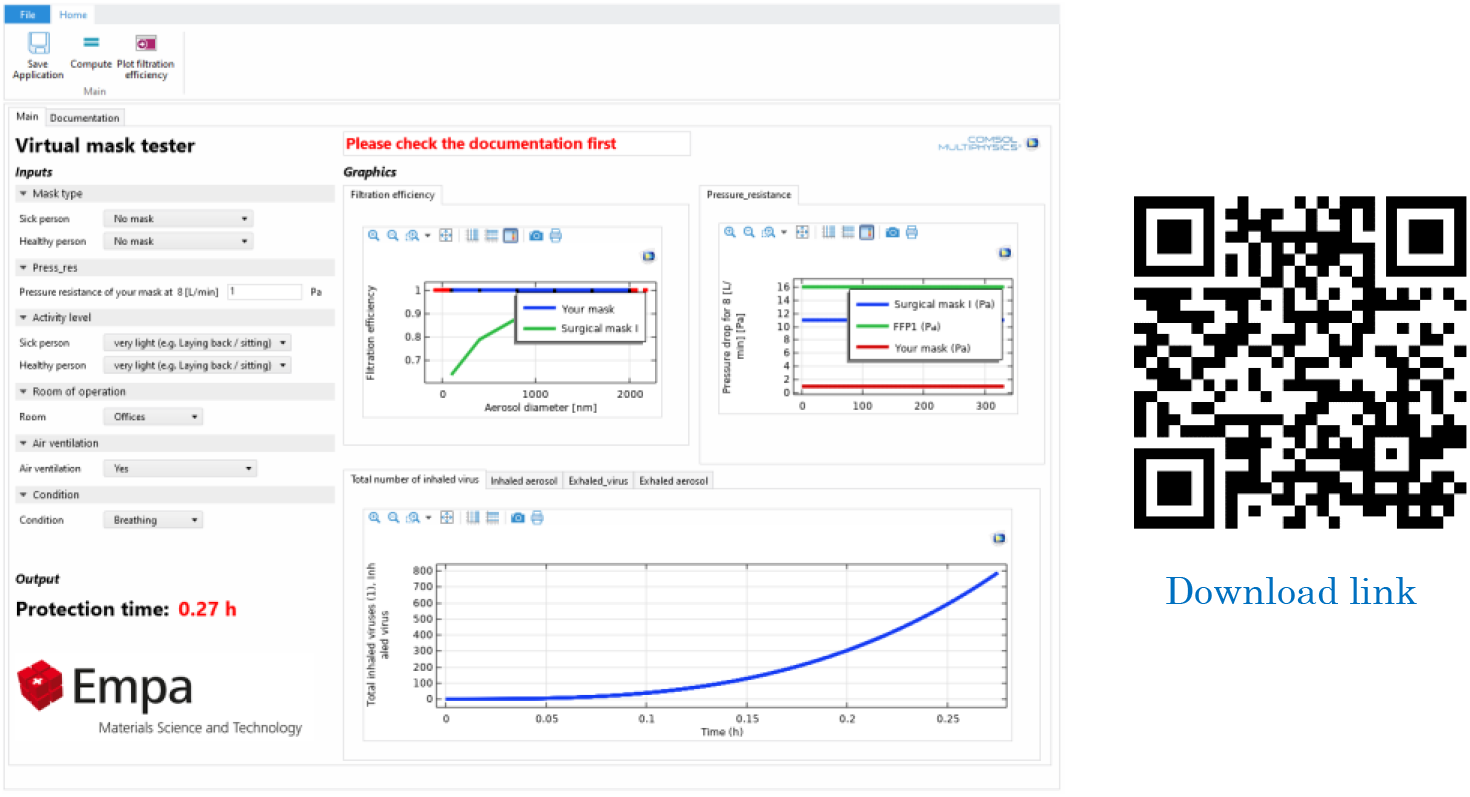
Virtual mask tester application interface.

## 5 CONCLUSIONS

Facemasks play an essential role in limiting the number of exhaled virus-laden aerosols in the environment, decreasing the risk of infection for healthy people, and reducing the risk of a pandemic. Based on our results, when the infected person wears a fully sealed FFP2, the number of emitted aerosols and viruses was reduced to 1/600 and 1/3500, respectively. Therefore, it clearly increases the protection time for the wearer. The difference observed in reducing the number of aerosols and viruses implies that besides the overall filtration efficiency, it is important to consider filtration of particular particle sizes to be highly relevant for virus transmission. Additional to the facemasks, the activity level of the infected person affected the emitted aerosols drastically. Based on the result, as the activity level of the infected person increases, the total emitted aerosols in the environment might increase up to 2500 times. This increase in the number of emitted aerosols affects the protection time, which is 300 times higher for the very low activity level than the very high activity level. This drastic change via activity shows the importance of additional safety measures in environments with highly active people. In the last step, we analyzed the effect of leakage on mask performance. Three types of masks were analyzed: surgical mask IIR, community mask, and FFP2. Based on the experimental input, the drop in the apparent filtration efficiency for surgical mask IIR is more drastic than the other two, leading to lower protection time. This result revealed that, despite the high filtration efficiency of the surgical mask, the leakage could diminish its performance. Besides the surgical mask IIR, the leakage for the community mask and FFP2 was considerable as well and reduced the protection time considerably. This result showed that as much as the filtration efficiency of a mask is important, the fitting of the mask on the face can also play an important role. In addition, the result of this study shows the impact of hygienic measures such as wearing masks and ventilating the rooms on protecting the people at risk of infection and their relevance for different types of rooms. The use of such a physics-based model to quantify the protection time of the wearer can be instrumental in evaluating new mask designs.

## Data Availability

All data produced in the present study are available upon reasonable request to the authors

## Acknowledgments

This work was supported by Innosuisse (ReMask research project, contract number 46668.1IP-EN). The authors declare that this study received funding from Innosuisse. The funder was not involved in the study design, collection, analysis, interpretation of data, the writing of this article, or the decision to submit it for publication. We would also like to thank Riccardo Innocenti Malini for sharing his insights at the beginning of the project on existing prior works.

